# Common variant approaches to study Mendelian disease gene function identify novel phenome and pathways associated with *PLOD3*

**DOI:** 10.1101/2025.11.25.25340832

**Authors:** Alexandra Scalici, James T. Baker, Freida Blostein, Megan Shuey, Dharmendra Choudhary, Ela W. Knapik, David C. Samuels, Jennifer E. Below, Lisa Bastarache, Tyne W. Miller-Fleming, Nancy J. Cox

## Abstract

**Background:** The study of rare and common genetic disorders, in terms of study design, methods, and their genetic architecture, has largely been thought of as distinct. As sequencing technologies and analysis methods have advanced, we have learned that polygenic background can affect the penetrance, severity, and onset of certain Mendelian conditions. While genome-wide analyses have significantly contributed to our understanding of human disease, these studies rarely explore how common variation within a traditionally Mendelian disease genes affects phenotypic outcomes.

**Methods:** In this study, we leverage common variant-based approaches and electronic health record (EHR)-derived phenome to study the phenotypic consequences associated with *PLOD3*, a Mendelian disease gene associated with BCARD syndrome. We conducted a gene-based phenome-wide association study (PheWAS) to identify phenotypes associated with reduced genetically predicted gene expression (GPGE) of *PLOD3* in BioVU. To further quantify the phenotypic features associated with *PLOD3*, we leveraged a phenotype risk score (PheRS) constructed from the Mendelian features BCARD syndrome in OMIM and a PheRS constructed from our gene-based PheWAS. We used these PheRSs in a TWAS farmwork to expand genetic and pathway level associations.

**Results:** We found that reduced GPGE of *PLOD3* derived from common variants, can capture clinical phenome associated with Mendelian disease (BCARD syndrome) in addition to novel phenotypes not previously associated with *PLOD3*. These novel phenotypes were largely replicated in analyses of a protein quantitative trait loci (pQTL) for PLOD3 and in identity by descent (IBD) analysis of the *PLOD3* locus. By generating PheRS for the Mendelian and gene-based phenome and using them in a TWAS framework, we identified novel genetic and pathway level associations.

**Conclusions:** In this study, we present a scalable approach using EHR-based phenome and GPGE to identify novel phenome and use it as a tool for gene discovery to identify novel gene and pathway associations. This study leveraged biobank scale genetic and phenotype data to identify candidate phenotypes for expansion of the *PLOD3* phenome and identify potential novel disease mechanisms. The application of these approaches to other Mendelian disease genes has the potential to aid in drug repurposing and identify candidate therapeutic targets.

## Introduction

We often classify the genetic architecture into two categories: Mendelian conditions associated with rare, high effect size genetic variation that are studied in family-based studies, and common disease associated with the small additive effects of common genetic variation from genome-wide association studies (GWAS).^1^ However, as the sample sizes of sequencing studies have increased, so has our ability to determine the effects of both common and rare variation on a wide range of complex phenotypes and to characterize the complex interplay between common and rare variation.

Recent studies have shed light on how polygenic background contributes to the phenome associated with Mendelian disorders often associated with rare, monogenic variation.^2,3^ For example, Huntington’s disease is an autosomal dominant Mendelian disorder caused by a CAG repeat expansion in the *HTT* gene and is diagnosed by genetic testing.^4^ Despite Huntington’s disease being classically Mendelian in nature, GWAS have identified common variation in the *HTT* region as well as other genes that are associated with the age of onset of Huntington’s.^5^ Other studies have deployed polygenic scores (PGS) to assess how the polygenic background can modify the penetrance by illustrating a risk gradient for Mendelian forms of coronary artery disease, colorectal cancer, breast cancer, and glaucoma.^6,7^ Expanding the genetic associations with classically Mendelian syndromes has the potential to enhance our understanding of disease mechanisms and expand potential therapeutic targets. Additionally, re-analysis of whole exome sequencing from study of families with known Mendelian syndromes and underlying genetic causes revealed that in 19 families where expansion of the known clinical phenome had been reported revealed that a large proportion of the expanded phenome was attributed to variation in other genes, illustrating the key role of multi-locus variation in the manifestation of the complex clinical phenotypes associated with Mendelian disorders.^8^ While these studies have shown that polygenic background can have effects in Mendelian conditions, there are limited studies on how common regulatory variation in Mendelian disease genes may impact phenotypic outcomes independent of coding sequence variation.

Vanderbilt University Medical Center’s extensive electronic health record (EHR) system contains approximately 3.5 million individuals with approximately 330,000 individuals with DNA samples and extensive phenotype data. Having access to BioVU, the EHR-linked DNA biorepository provides access to both genotype and phenotype information for a large quantity of individuals.^9,10^ Leveraging EHR-linked genetic information in biobanks such as BioVU has led to the development of methods allowing for rare Mendelian diseases to be studied at a large scale. The implementation of phenotype risk scores (PheRS), which leverage EHR-derived phenotypes, has been instrumental in the identification of individuals with undiagnosed rare disease, and contributed to our understanding of the broad range of phenome associated with Mendelian disease.^11,12^ Studies have also integrated genetically predicted gene expression (GPGE) from transcriptome-wide association study (TWAS) methods derived from common variation captured on genotyping arrays to identify medical phenome associated with reduced GPGE of Mendelian disease genes in BioVU. For example, a study of reduced GPGE of *CFTR* was able to capture phenotypes consistent with the clinical diagnosis of cystic fibrosis.^13^ By examining the phenome associated with reduced GPGE of *RIC1* in conjunction with zebrafish models and patient data, this study was able to more fully characterize the phenome of a *RIC1*-associated Mendelian disorder, CATIFA syndrome (cleft lip, cataracts, tooth abnormality, impaired intellectual development, facial dysmorphism, and attention-deficit hyperactivity disorder (ADHD) (MIM # 618761)).^14^ In addition to examining the relationship between phenome and GPGE, we used DRIVE, a tool that identifies networks of individuals sharing pairwise IBD segments around a target locus and assesses phenotypic enrichment within these networks.^15^ These BioVU based studies illustrate the power of integrating EHR-derived phenotypes with genetic data in aiding in our understanding of genetic conditions.

BCARD syndrome (Lysyl Hydroxylase-3 (LH3) deficiency) (MIM# 612394) is a Mendelian disorder phenotypically characterized by bone abnormalities with joint contractures, cataracts, arterial rupture, and deafness (BCARD) and has been associated with rare variants in *PLOD3.*^16–19^ *PLOD3* encodes procollagen-lysine,2-oxoglutarate 5-dioxygenase 3, which is responsible for post-translational modifications to collagen fibrils within the endoplasmic reticulum that are essential for proper collagen crosslinking in the extracellular matrix (ECM).^20–22^ Additional clinical features such as bullous dermatoses and diaphragmatic eventration, focal epilepsy, intestinal abnormalities, and altered brain morphology have also been reported.^17,19,23^ BCARD syndrome is very rare, with an estimated prevalence of approximately 1 in 1 million [ORPHA:300284] individuals globally and only 7 cases have been reported in OMIM; therefore, studies of this disorder have been limited to case reports and family-based studies.^24^ While these case reports and family studies focus on rare variation in *PLOD3*, a GWAS of blood serum protein levels identified a protein quantitative trait locus (pQTL) for PLOD3 serum levels.^25^

Here we present a scalable approach to better understand the underlying genetic mechanisms driving the broad spectrum of clinical phenotypes that characterize Mendelian disorders. We applied this analytic framework to *PLOD3,* a gene with rare variation that has been associated with BCARD syndrome. We leveraged common variation captured on genotyping arrays within BioVU, VUMC’s EHR-linked biobank, to conduct gene and phenome-based analyses. We implemented several TWAS approaches to obtain GPGE of *PLOD3* and conducted a PheWAS to identify medical phenome associated with reduced GPGE of *PLOD3.* We used the phenome identified in this analysis and the Mendelian features of BCARD syndrome listed on OMIM to construct two PheRSs. To better understand the phenome potentially being driven by rare variation in *PLOD3*, we implemented identity by descent (IBD) based approaches to examine genetic sharing across unrelated individuals at the *PLOD3* locus and phenotypic enrichment among those with IBD sharing. We used both this gene-based PheRS and the Mendelian PheRS in a TWAS to expand gene and pathway associations to provide additional insights into the underlying mechanisms driving the phenome observed in BCARD syndrome.

## Materials and Methods

### Vanderbilt University Medical Center’s electronic health record system and BioVU

VUMC has an extensive de-identified health record (EHR) system used for biomedical research known as the synthetic derivative (SD). The SD contains over 3.5 million individuals with various health information available such as ICD9 and ICD10 billing codes, procedure codes, clinical notes, and laboratory values.^9,10,26^ BioVU, VUMC’s EHR-linked biobank contains over 330,000 individuals with DNA samples. Of these, we used 70,439 individuals of genetically determined European ancestry (determined by principal component analysis (PCA)) that were genotyped using the Illumina MEGA^Ex^ array. The genetic data has passed rigorous quality control standards as previously described.^26^ All analyses of *PLOD3* were conducted using these 70,439 BioVU participants of European ancestry.

### *PLOD3* gene-based phenome-wide association study (PheWAS)

We have implemented machine learning models such as PrediXcan, unified test for molecular signatures (UTMOST), and Joint Tissue Imputation (JTI) to calculate GPGE for all 70, 439 BioVU participants of European ancestry^27–30^. Using the best performing prediction model (JTI) and the best performing gene-tissue pair for PLOD3 GPGE (Kidney Cortex, r^2^ = 0.10), we conducted a gene-based PheWAS to test the association between 1817 phecodes and *PLOD3* GPGE adjusting for age, sex, the first 10 principal components of ancestry, and number of visits using the PheWAS R package.^13,14,31,32^ We required a minimum number of cases for each phecode (n>20) to be included in the analyses. The direction of effect (beta) indicates whether each phenotype is associated with increased or reduced GPGE. The implementation of a gene-based PheWAS allows us to identify medical phenome associated with reduced GPGE of *PLOD3* and compare it to the established clinical features of BCARD syndrome.

### Laboratory value association study with reduced *PLOD3* GPGE

Our PheWAS found that both hypothyroidism and Grave’s disease, an autoimmune disorder associated with hyperthyroidism, were associated with reduced GPGE of *PLOD3.* To further explore the relationship between *PLOD3* and thyroid dysfunction, we tested whether reduced GPGE of *PLOD3* is associated with thyroid-stimulating hormone (TSH), which is often used to diagnose hypo- and hyperthyroidism. Laboratory values have undergone rigorous quality control through the quality labs pipeline.^26,33^ To test the association between *PLOD3* GPGE and hypothyroidism, we tested whether *PLOD3* GPGE is predictive of median age-adjusted TSH levels using age, sex, the first 10 principal components of ancestry, and number of visits as covariates in a linear regression model.

### *PLOD3* gene-based PheRS construction and application

Using the beta estimates from the phenome that is significantly enriched among participants with reduced *PLOD3* GPGE (p<0.05) from our gene-based PheWAS, we constructed a *PLOD3* PheRS^11,12,34^ (**Table S5**). With this weighting scheme, individuals with a negative PheRS have more of the phenome associated with reduced GPGE of *PLOD3*. We used this PheRS in a TWAS to identify genes with altered GPGE associated with the *PLOD3* phenome. Using the significantly associated genes from our TWAS (p<0.05), we conducted an over-representation analysis (ORA) using WebGestalt.^35,36^ Of the 1,395 genes uploaded to WebGestalt, 1,106 mapped to EntrezGene IDs. 357 of these genes were annotated to functional categories within the KEGG pathway database setting the minimum number of genes within each category to 5 and a false discovery rate (FDR<5%).

### Mendelian PheRS construction and application

BCARD syndrome is one of over 2,000 Mendelian disorders that had a PheRS constructed within BioVU using the clinical features described in the Online Mendelian Inheritance of Man (OMIM) and a log-transformed inverse prevalence weighting system as previously described^11,12^ (**Table S6**). Using this weighting scheme, a higher PheRS indicates that an individual has more of the BCARD phenotypes. We used this BCARD syndrome PheRS in a TWAS to identify changes in GPGE associated with the Mendelian disease phenome. Using the significantly associated genes from our TWAS (p<0.05), we conducted over-representation analysis (ORA) we conducted an over representation analysis (ORA) using WebGestalt.^35,36^ Of the 1390 genes uploaded to WebGestalt, 971 mapped to entrezgene IDs. 299 of these genes were annotated to functional categories within the KEGG pathway database setting the minimum number of genes within each category to 5 and a false discovery rate (FDR<5%).

### *PLOD3* pQTL phenome-wide association study (PheWAS)

We searched the NHGRI EBI-GWAS Catalogue for SNP associations that were identified in the *PLOD3* locus.^37^ We found that one SNP (rs41281013) where the alternate allele (G) is associated with reduced blood serum levels of PLOD3 protein.^25^ To identify medical phenome that is associated with this SNP, we conducted a PheWAS using the PheWAS R Package.^32,34,38^ We ran a logistic regression model testing the association of each phecode with SNP dosage of the alternate allele coded as 0,1,2, adjusting for age, sex, the first 10 principal components of ancestry, and the number of visits with a Bonferroni threshold of p-value < 2.94 x 10^-5^.

### Calculating identity by descent (IBD) sharing at the *PLOD3* locus using Distant Relatedness for Identification and Variant Evaluation (DRIVE) & phenome-wide enrichment study (PheWES)

Pairwise IBD segments were first identified within the cohort of European ancestry in BioVU using hap-IBD.^39^ To assess whether individuals of European ancestry in BioVU carried shared haplotype segments at the *PLOD3* locus (chr7:100849266-100860862, GRCh37), we used the DRIVE tool.^15^ DRIVE was run with default parameters, with a minimum segment threshold set to 3cM. Using these networks of individuals with IBD sharing at the *PLOD3* locus, we conducted a PheWES to determine the enrichment of medical phenome among individuals within a network compared to the total cohort.

## Results

### PheWAS of reduced GPGE of *PLOD3* captures phenome associated with rare variation in *PLOD3* as well as phenome not previously associated with rare variation in *PLOD3*

Rare variation in *PLOD3* has been associated with BCARD syndrome which clinically presents with bone fragility, contracture of joints, cataracts, arterial rupture, deafness, diaphragmatic extraversion, developmental delay, and skin blistering (MIM **#** 612394)^16,17,19^. We found that among the range of phenotypes significantly associated with reduced *PLOD3* GPGE (p<0.05) including contracture of joints (OR=0.87, p= 0.004), acquired deformities of finger (OR=0.82, p=0.001), non-healing surgical wound (OR=0.78,p=0.0004), diaphragmatic hernia (OR=0.94, p=0.013), hemorrhagic disorder due to intrinsic circulating anticoagulants (OR=0.85, p=0.02), and learning disorder (OR=0.75, p=0.03) (**Figure 1**, **Table 1, Table S1**).

**Figure 1:**
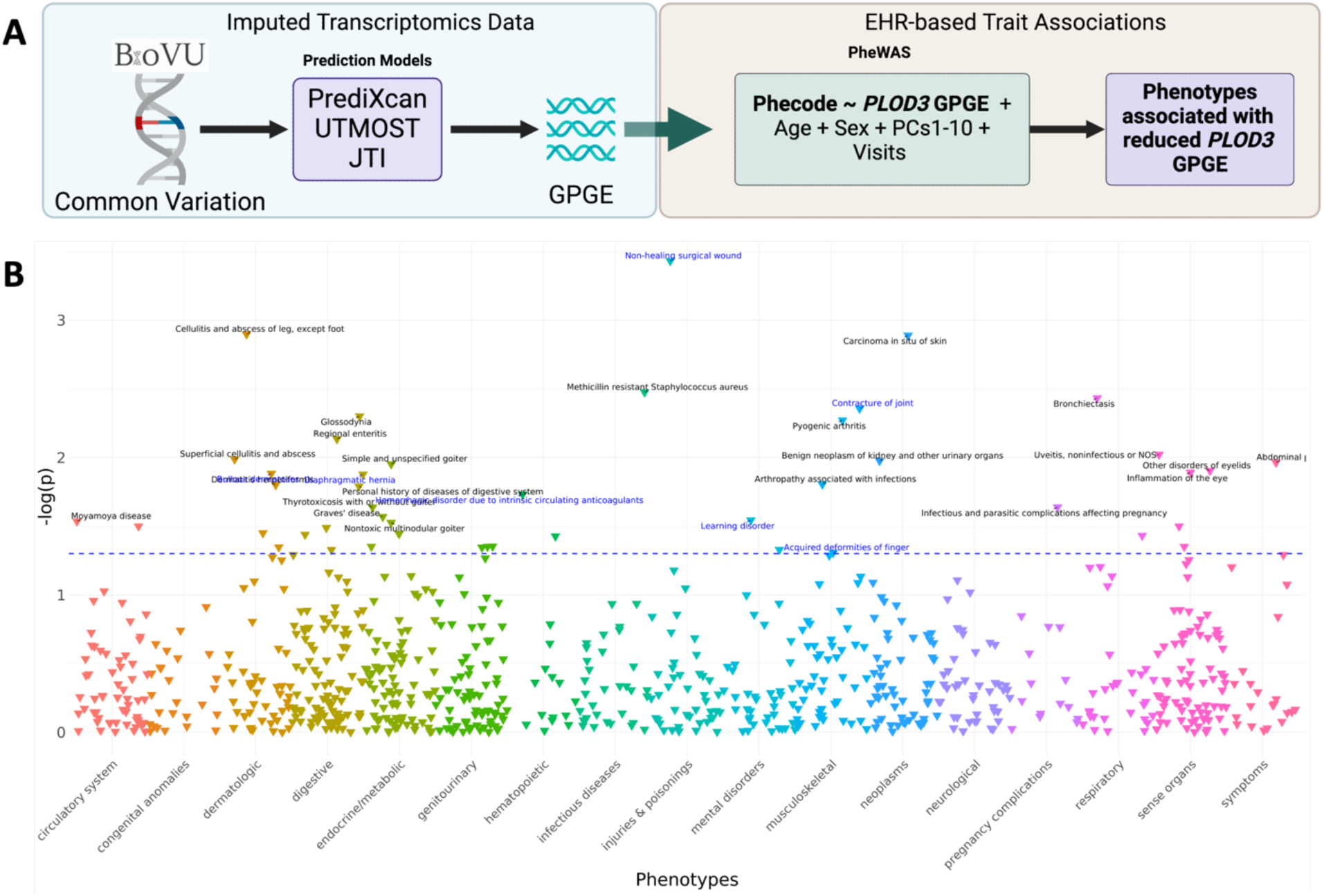
PheWAS of reduced GPGE of *PLOD3* capture phenome associated with rare variation in *PLOD3* as well as phenome not previously associated with rare variation in *PLOD3*. A) A diagram depicting an analysis overview*. **B)** Phenome associated with reduced GPGE of *PLOD3.* The blue dotted line represents a significance level of 0.05 *Created with biorender.com

**Table 1:**
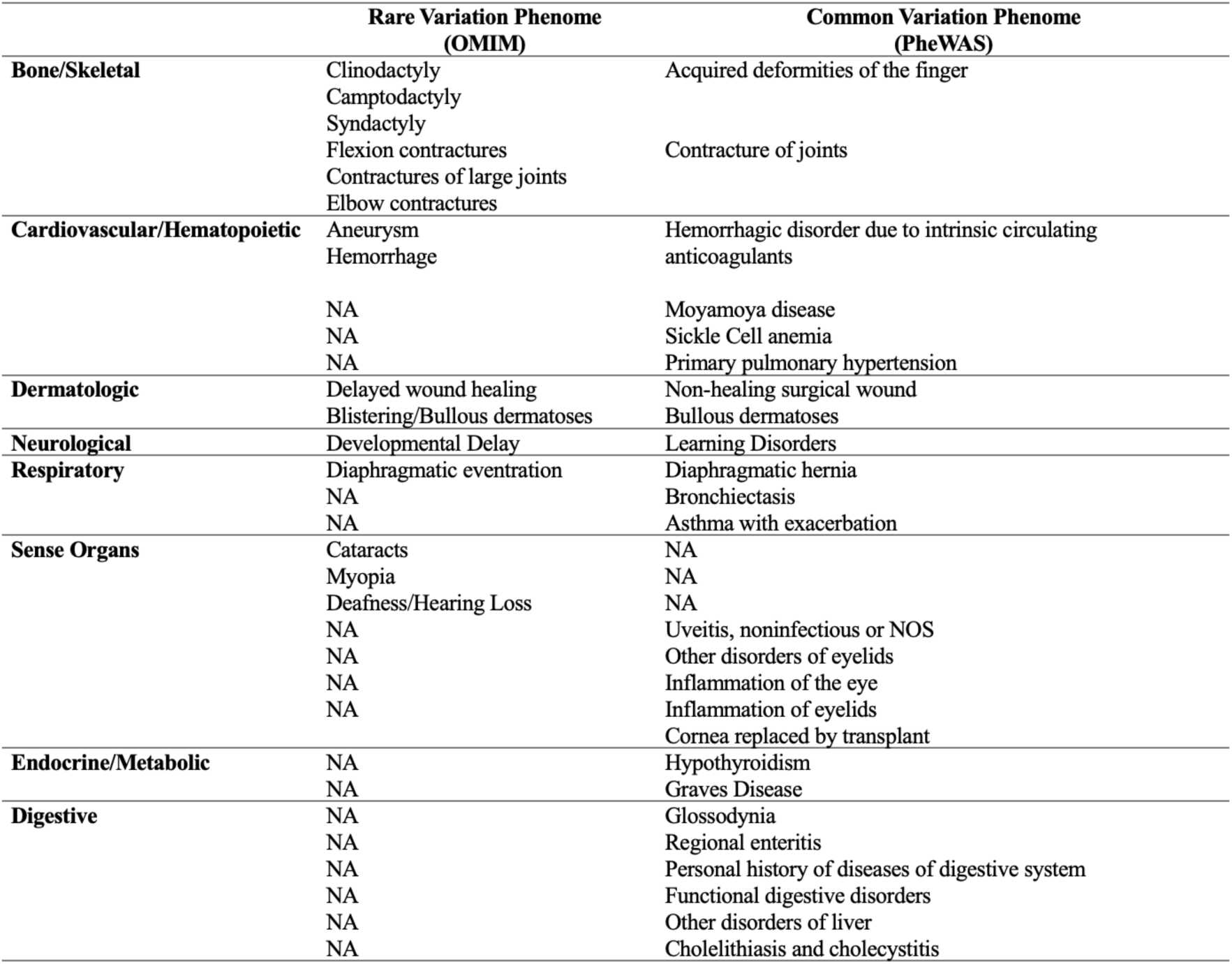
BCARD and *PLOD3-*associated phenome.

### Reduced GPGE of *PLOD3* is associated with increased levels of TSH

Phenotypes such as hypothyroidism have not been associated with variation in *PLOD3* and human disease. To further investigate the role of *PLOD3* in hypothyroidism, we tested whether *PLOD3* GPGE is associated with normalized laboratory values of thyroid-stimulating hormone (TSH). Among the individuals of European ancestry in BioVU (N = 70,439), we had 27,247 individuals with TSH labs.

Using this subset, we tested the association of *PLOD3* GPGE with TSH lab values in a linear regression model adjusting for age, sex, the first ten principal components of ancestry, and number of visits. We found that for every 1 standard deviation unit increase in *PLOD3* GPGE, TSH levels decrease by 0.0141 units (β=-0.0141, p=2.2×10^-2^), indicating that reduced *PLOD3* GPGE is correlated with increased TSH levels, which is often an indicator of hypothyroidism (**Table S2**).

### PheWES of networks of IBD sharing at *PLOD3* identifies core medical phenome associated with rare and common variation in *PLOD3*

DRIVE identified 12,295 networks of individuals among the BioVU participants of European ancestry who shared pairwise IBD segments containing the *PLOD3* locus (**Table S3**). By conducting a PheWES analysis across these networks, we identified networks with IBD sharing of *PLOD3* that were enriched for musculoskeletal phenotypes such as synovitis and tenosynovitis (p=3.06e-05), spondylosis and allied disorders (p=0.00013318), muscle weakness (p=0.00014469), joint/ligament sprain (p= 0.000522704), acquired foot deformities (p=0.00024204), chondrocalcinosis (p=0.00071773), acquired toe deformities (p=0.0007553), and Intervertebral Disc Disease (IVDD) (p=0.00079499), hemorrhage phenotypes such as hemorrhage of rectum and anus (p= 0.000312147), gastrointestinal hemorrhage, (p= 0.000926698), hemorrhage or hematoma complicating a procedure (p= 0.001378605), intracranial hemorrhage (p= 0.003558836), eye phenotypes such as visual disturbances (p=0.00025919), dermatologic phenotypes such as degenerative skin conditions and other dermatoses (p=0.000287), and respiratory phenotypes such as diaphragmatic hernia (p=8.24e-06) (**Table S4**). These phenotypes represent some of the critical hallmark features that have been clinically described in BCARD syndrome.

In addition to these hallmark phenotypes, new clinical studies have identified novel variants in *PLOD3* among BCARD patients that have been associated with novel phenome such as digestive abnormalities and expanded brain phenotypes. Our PheWES analysis of IBD sharing of *PLOD3* identified networks enriched for intestinal abnormalities such as ulceration of intestine (p= 1.18e-05) and diverticulitis (p= 0.000183236) and neurological phenotypes such as psychogenic and somatoform disorders (p= 0.000208879), memory loss (p= 0.000209124), altered mental status (p= 0.00037156), insomnia (p= 0.000399896), major depressive disorder (p= 0.000481438), and dementia (p= 0.000551207) (**Table S4**).

### Individuals with longer shared segment lengths at the *PLOD3* locus tend to have lower differences in both their Mendelian and gene-based PheRS

To compare shared segment length and shared *PLOD3* phenome, we calculated the pairwise shared IBD segment length within the *PLOD3* locus and the absolute value of the difference of their Mendelian and gene-based PheRS. When comparing the length of each shared segment with the absolute value of the PheRS difference, we observed that as shared segment length increases, the differences in PheRS are minimal (**Figure S1**). This indicates that individuals with greater IBD sharing at the *PLOD3* locus, also share similar phenome.

### Gene-based PheRS TWAS identifies distinct gene and pathway-level associations

Using the PheRS constructed from our PheWAS of *PLOD3* GPGE in a TWAS, we identified several genes associated with the *PLOD3* phenome. Eight genes were associated with the *PLOD3* PheRS including *PLOD3* (β = 0.005, p = 4.83 x 10^-52^), *PILRA* (β = 0.001, p = 2.25 x 10^-6^), *AC092849.1* (β = -0.001, p = 4.36 x 10^-6^), *PILRB* (β = 0.001, p = 9.96 x 10^-6^), *SAP25* (β = 0.001, p = 9.96 x 10^-5^), *MYL10* (β = -0.001, p = 4.90 x 10^-5^), *TSC22D4* (β = 0.001, p = 7.61 x 10^-5^), and *HIST1H1E* (β = -0.001, p = 1.73 x 10^-5^) (**Figure 2A-B, Table S7)**. This analysis seems to identify a large region on chromosome 7 surrounding PLOD3, indicating that these associations may not be independent. Using the nominally significant gene associations (p<0.05) with our *PLOD3* PheRS, we conducted an overrepresentation analysis (ORA) using WebGestalt. We found an overrepresentation of pathways among our gene set such as pyruvate metabolism (p=0.002), hypertrophic cardiomyopathy (p=0.006), notch signaling pathway (p=0.007), glycolysis/gluconeogenesis (p=0.02), propanoate metabolism (p=0.02), base excision repair (p=0.02), cysteine and methionine metabolism (p=0.02), glucagon signaling (p=0.02), dilated cardiomyopathy (p=0.03), PI3K-Akt signaling pathway (p=0.05) (**Figure 2C, Table S8**)

**Figure 2:**
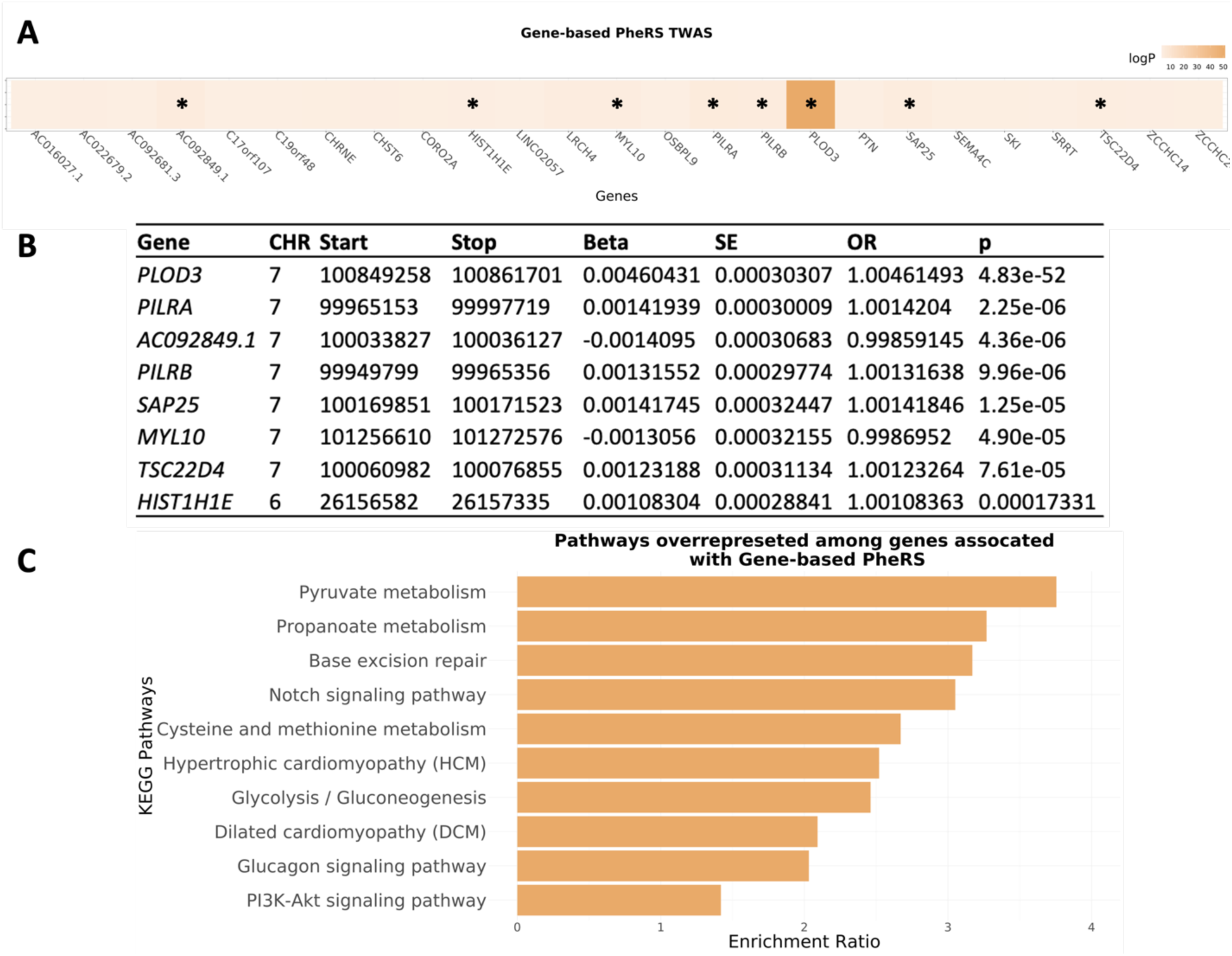
Genes and pathways associated with the *PLOD3* PheRS. **A)** A heat map of the top 25 most significant GPGE associations with the *PLOD3* PheRS. The asterisk (*) represents genes that passed a Bonferroni correction. **B)** A table of the Bonferroni significant genes. **C)** Pathways overrepresented among nominally significant genes associated with *PLOD3* PheRS (p<0.05).

### Mendelian PheRS TWAS identifies novel gene and pathway-level associations

The BCARD syndrome phenome described in OMIM was mapped to HPO terms and subsequently phecodes to construct a Mendelian PheRS comprised of sixteen phecodes using a log-transformed inverse prevalence weighting scheme in BioVU.^11^ (**Table S6)** We used this BCARD syndrome PheRS in a TWAS framework to expand gene and pathway-level associations. Two genes that passed our multiple testing correction: *AC053503.4* (β = 0.01575917, p = 5.35E-05) and *AC005838.2* (β = -0.0143819, p = 6.35E-05) (**Figure 3A-B**). Both are long non-coding RNAs (lncRNAs) with unknown targets. Additionally, reduced GPGE of *PLOD3* is associated with this Mendelian PheRS (β = -0.0082607, p = 0.02486628), indicating that this PheRS is capturing phenome related to *PLOD3* and its associated biology (**Table S9**).

**Figure 3:**
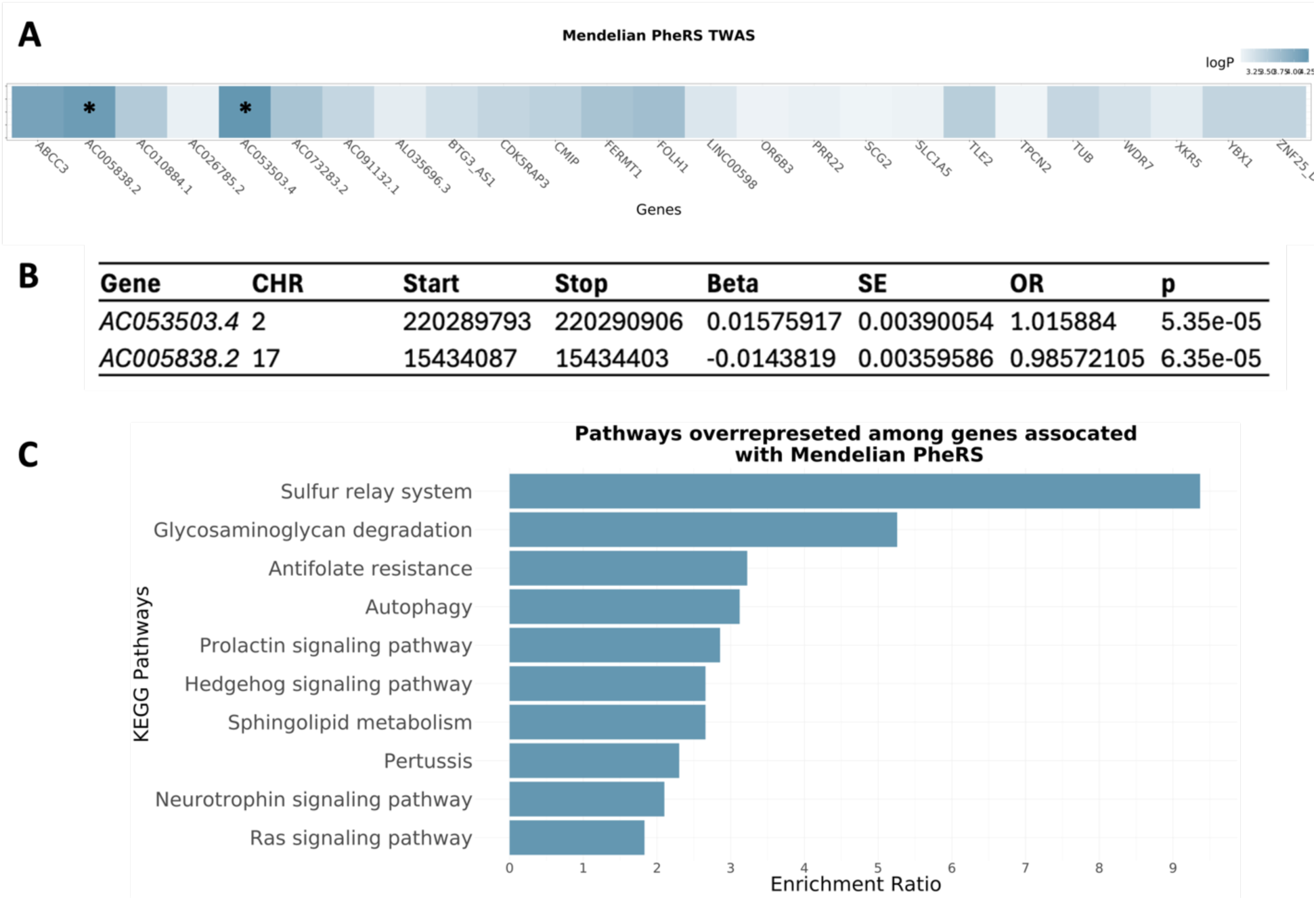
Genes and pathways associated with the BCARD PheRS. **A)** A heat map of the top 25 most significant GPGE associations with the BCARD PheRS. The asterisk (*) represents genes that passed a Bonferroni correction. **B)** A table of the Bonferroni significant genes. **C)** Pathways overrepresented among nominally significant genes associated with BCARD PheRS (p<0.05).

Using the nominally significant PheRS TWAS associations (p<0.05), we conducted an overrepresentation analysis (ORA) using KEGG pathways in WebGestalt. We identified an over representation of genes in pathways such as the sulfur relay system (p=0.003), glycosaminoglycan degradation (p=0.006), antifolate resistance (p=0.03), autophagy (p=0.04), prolactin signaling (p=0.007), hedgehog signaling (p=0.04), sphingolipid metabolism (p=0.04), Pertussis (p=0.38), neurotrophins signaling pathway (p=0.02), and Ras signaling pathway (p=0.01) (**Figure 3C, Table S10).**

### *PLOD3* pQTL PheWAS illustrates that common variation in *PLOD3* is associated with connective tissue and musculoskeletal defects

By examining the phenome associated with the pQTL for PLOD3 serum protein levels listed in the GWAS catalog (**Figure 4A)**, we found that skeletal defects such as congenital anomalies of the face and skull (β = 0.78552219, p = 0.039), other congenital musculoskeletal anomalies (β = 0.78200601, p = 0.005), congenital osteodystrophies (β = 1.56324633, p = 0.00059339), osteoarthrosis (β = -0.29248, p = 0.0096012), fracture of vertebral column without mention of spinal cord injury (β = 0.39303199, p = 0.0345054), and fracture of tibia and fibula (β = 0.54217691, p = 0.54217691) (**Figure 4B, Table S11**). These results capture some of the key musculoskeletal features of BCARD syndrome such as bone fragility and craniofacial anomalies captured in OMIM.

**Figure 4:**
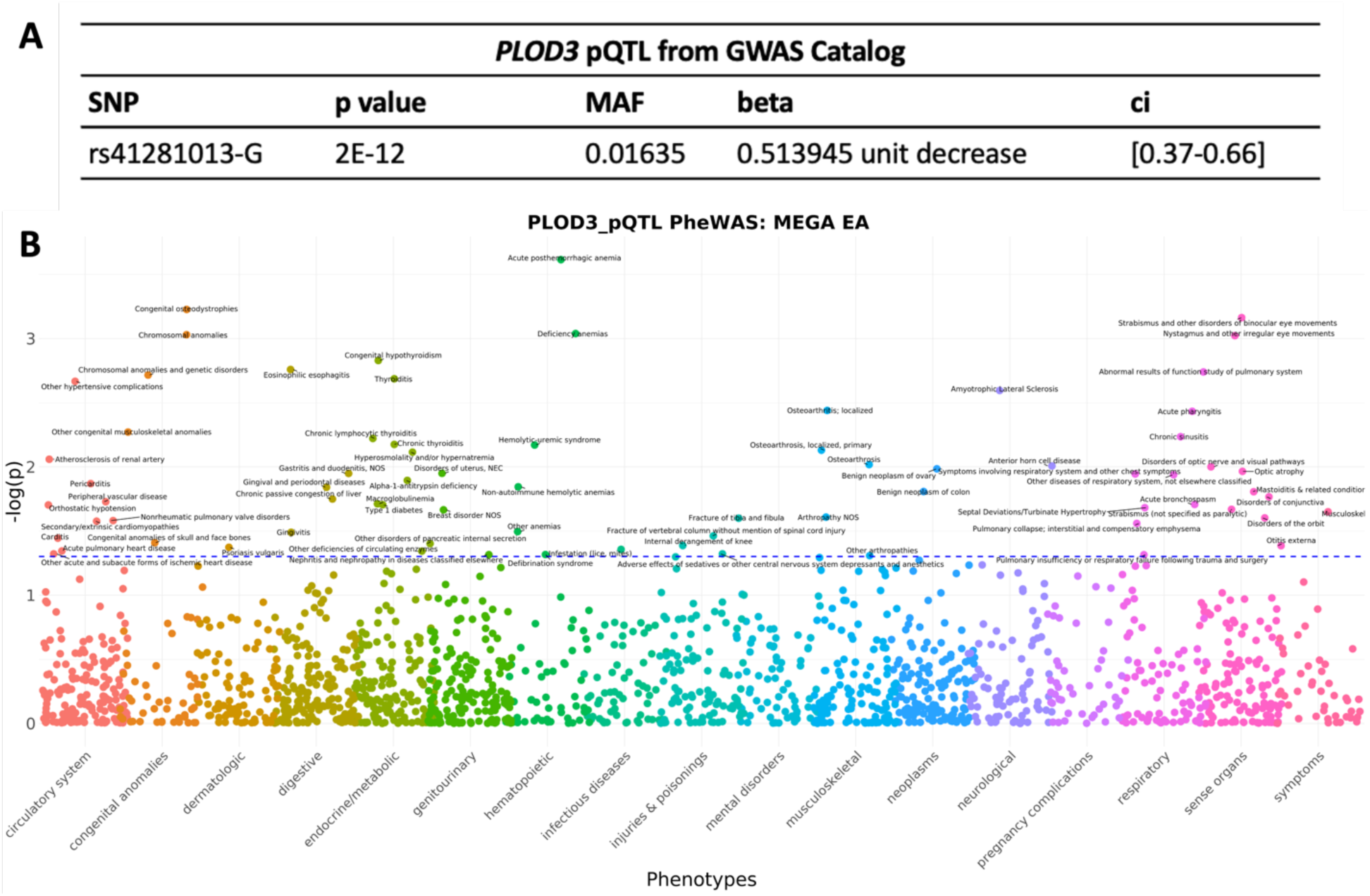
PLOD3 pQTL captures Mendelian and novel phenome. **A)** pQTL variant information from the GWAS catalog. **B)** Plot illustrating the phenotype associations with the PLOD3 pQTL. The dotted line indicates a significance level of p =0.05.

### *PLOD3* pQTL PheWAS illustrates that common variation in *PLOD3* is associated with novel phenotypes captured in our gene-based methods

In addition to these key musculoskeletal features such as congenital anomalies of the face and skull, other congenital musculoskeletal anomalies, congenital osteodystrophies, osteoarthrosis, fracture of vertebral column without mention of spinal cord injury, and fracture of tibia and fibula, that are a hallmark of BCARD syndrome, the phenome associated with this pQTL also captures phenotypes such as thyroid dysfunction (congenital hypothyroidism (β = 1.33769536, p = 0.00148312), thyroiditis (β = 0.60878083, p = 0.00205747)) and anemias (deficiency anemias (β = 1.09600677, p = 0.00091429), non-autoimmune hemolytic anemias (β = 1.23656803, p = 0.01430211), other anemias (β = 0.17821355, p = 0.03195327) and acute posthemorrhagic anemia (β = 0.41577625, p = 0.00024276)), further supporting the hypothesis that these phenotypes are driven by common variation in *PLOD3* (**Figure 4B, Table S11**).

## Discussion

The use of gene-based PheWAS has allowed us to gain a greater understanding of how common variation in a Mendelian disease gene affect phenotypic outcomes and our understanding of basic gene function despite not having access to individuals diagnosed with BCARD syndrome due to its low prevalence. Conducting a gene-based PheWAS examining the medical phenome associated with reduced GPGE of Mendelian disease genes is an established method for capturing the phenome associated with Mendelian disease and identifying candidate phenotypes for phenotypic expansion.^13,14^ By applying this gene-based PheWAS method to identify phenome associated with reduced GPGE of *PLOD3*, we found that we are able to capture some of the key features of BCARD syndrome, such as musculoskeletal defects (contracture of the joints, acquired deformities of the finger), dermatologic phenotypes (bullous dermatoses), diaphragmatic abnormalities, and hemorrhage (**Figure 1B**, **Table 1**). Our ability to capture specific features of the Mendelian disorder described in OMIM, further illustrates that using reduced GPGE can model the phenotypic consequences of rare loss-of-function variants in a Mendelian disease gene. Additionally, we leveraged the DRIVE tool to identify shared IBD segments containing the *PLOD3* locus. Because these shared IBD segments are more likely to harbor rare variation, we would expect that unrelated individuals that share segments IBD would have shared phenome, and more specifically, share phenome that has been associated with rare variation in *PLOD3*. Although BioVU lacks individuals affected by BCARD syndrome, our PheWES analysis identified some of the core phenotypes associated with rare variation in *PLOD3* and BCARD syndrome. The trend between having longer segment length and less of a difference in both the Mendelian and gene-based PheRS provides a means to quantify and examine patterns of genetic and phenotypic sharing associated with a single gene.

In addition to capturing some of the Mendelian features of BCARD syndrome, our gene-based PheWAS also identified phenotypes that have not been previously associated with variation in *PLOD3* such as disorders of the thyroid (Hypothyroidism, Graves’ Disease, and increased TSH laboratory values) and hematopoietic phenotypes (Sickle Cell Anemia and Moyamoya disease) (**Table 1**). While *PLOD3* has not previously been associated with thyroid dysfunction in humans, *Plod3* expression was found to be a biomarker for hypothyroidism in rats.^40^ Having evidence suggesting that *PLOD3* may play a role in thyroid dysfunction in both our biobank analysis and in a rat model identifies the potential candidate phenotypes for phenotypic expansion and suggests that further investigation of thyroid disease in patients with variants in *PLOD3* may be warranted. Our gene-based PheWAS, our IBD analysis, and our pQTL all identified significant anemia-related phenotypes. While there has been no direct link to of variation in *PLOD3* to different types of anemia, a clinical case report of an individual with BCARD syndrome reported pregnancy complications due to anemia.^23^ Additionally, iron is a cofactor for PLOD3, which aids in its primary catalytic function, the hydroxylation of lysyl residues during collagen synthesis.^41^ While further investigation is required, we suspect these strong associations with anemia related phenotypes is not a direct result of variation in *PLOD3*, but rather comorbid conditions that result from iron deficiency. This highlights the ability of these gene-based and common variant approaches to identify phenome that is related to the underlying genetic mechanisms, gene function, and phenotypic outcomes. While this gene-based PheWAS approach only identifies phenotypes at a nominal significance level, previous applications of this approach were able to identify phenotypes that were clinically validated in patients and functionally validated in zebrafish models.^13,14,31^ Scaling these methods to other Mendelian disease genes can aid in our understanding of how these complex multisystemic genetic syndromes clinically present and identify potential candidates for phenotypic expansion and allow us to gain a greater understanding of the genetic mechanisms driving the complex phenome associated with Mendelian disorders.

PheRSs have been successfully used to identify patients with undiagnosed genetic disease, by leveraging the unique combinations of phenotypes that often clinically present in patients with rare, Mendelian disorders at biobank scale.^11,12^ A recent study in the common disease space deployed PheRS as a tool for gene and pathway discovery to provide biological context to the shared genetic architecture of eye disease, providing an example of how using EHR-derived phenome can aid in expanding genetic and pathway-level associations.^42^ In this study, we leverage similar tools to gain greater understanding of the phenome and pathways associated with a single gene. We used both the BCARD PheRS and a gene-based *PLOD3* PheRS to expand genes and pathways associated with the *PLOD3* phenome in a TWAS. The genes identified from our PheRS TWAS were all located within the same region on chromosome 7, surrounding the *PLOD3* locus, indicating that these gene-level associations are not independent and likely confounded by linkage disequilibrium or complex regulatory architecture in the region. Our pathway analysis results for genes nominally associated with both PheRS revealed two interesting trends. First, we observed a strong enrichment of metabolic pathways such as glycosaminoglycan (GAG) degradation, sphingolipid metabolism, pyruvate metabolism propanoate metabolism, cysteine and methionine metabolism, glycolysis/gluconeogenesis and glucagon signaling pathway (**Figure 2C and Figure 3C**). This suggests that metabolic dysfunction may be a consequence of loss of or reduced GPGE of *PLOD3.* Second, pathways such as neurotrophin signaling, autophagy, RAS signaling, and PI3K-AKT signaling are all major regulators of MAPK signaling, playing a critical role in cell survival and death.^43–45^

Expanding the genetic pathways associated with Mendelian disease genes not only gives us insight into potential biological mechanisms but also can aid in expanding potential therapeutic targets. The hydrolysis of lysyl residues by *PLOD3* during collagen synthesis results in the production of hydroxylysine, succinate, and carbon dioxide as byproducts. In addition to being a byproduct of the catalytic activity of PLOD3, succinate is also a required element in the Krebs cycle.^46–48^ Additionally, the BROAD Institute Drug Repurposing Hub lists *PLOD3* as a potential target gene for succinic acid.^49^ This biochemical data previously reported in the literature in conjunction with our analyses identifying metabolic phenotypes and pathways associated with *PLOD3,* further suggests that reduced or loss of *PLOD3* can disrupt metabolic activity. Functional validation studies in loss of function zebrafish models of *plod3* are currently underway.

The goal of this study is to leverage common variation within a single gene to gain greater insight into gene function and potential mechanisms underlying human disease. By examining the medical phenome associated with reduced GPGE of *PLOD3* and phenotypic enrichment analysis in networks of unrelated individuals that share genetic segments IBD at the *PLOD3* locus, we can capture phenotypes associated with BCARD syndrome, a rare Mendelian disorder linked to rare variation in *PLOD3,* as well as novel phenotypes. Identification of these novel phenotypes put forth potential candidates for phenotypic expansion and highlights the ability to scale this method to other Mendelian disease genes to gain greater insights into clinically relevant phenome and disease mechanisms. Additionally, we further quantified this phenome into a PheRS and identified novel genes and pathways associated with the PheRS. Using PheRS as a tool for novel gene and pathway discovery in the context of human disease can aid in expanding therapeutic targets for rare genetic disorders. While further functional analyses are required to understand the true underlying mechanism driving the complex phenome associated with *PLOD3* and BCARD syndrome, our goal is to provide the foundation and starting point for these functional analyses.

## Author Contributions

**A.S** conducted formal analyses, code and pipeline development, drafted manuscript. **J.T.B** conducted formal analysis, reviewed, and edited manuscript., **F.B.** curated data sets, reviewed and edited manuscript., **M.M.S** interpreted results, reviewed and edited manuscript., **D.C.** contributed to study conceptualization, reviewed and edited manuscript. **E.W.K** contributed to study conceptualization, reviewed and edited manuscript., **D.C.S** interpreted results, reviewed and edited manuscript., **J.E.B.** interpreted results, reviewed and edited manuscript., **L.B.** interpreted results, reviewed and edited manuscript. **T.M.F.** curated data sets, code and pipeline development, interpreted results, reviewed and edited manuscript., **N.J.C** study conceptualization, reviewed and edited manuscript, and supervised the project.

## Data and code availability

Information on how to collaborate and the BioVU data sharing policy can be found at: https://victr.vumc.org/how-to-use-biovu/

All results obtained from the analyses described in this manuscript are included in the supplementary tables.

## Funding Sources

The Vanderbilt University Medical Center Synthetic Derivative, which is supported by institutional funding, the 1S10RR025141-01 instrumentation award, and the CTSA grant UL1TR000445 from the National Center for Advancing Translational Sciences/National Institutes of Health. A.S. was supported by the 5T32GM080178-15 and 1T32GM145734-01 from the Training Program in Genetic Variation & Human Phenotypes. M.M.S. was supported by the National Institutes of Health (K12HD043483).

## Supplemental Figures

**Figure S1:**
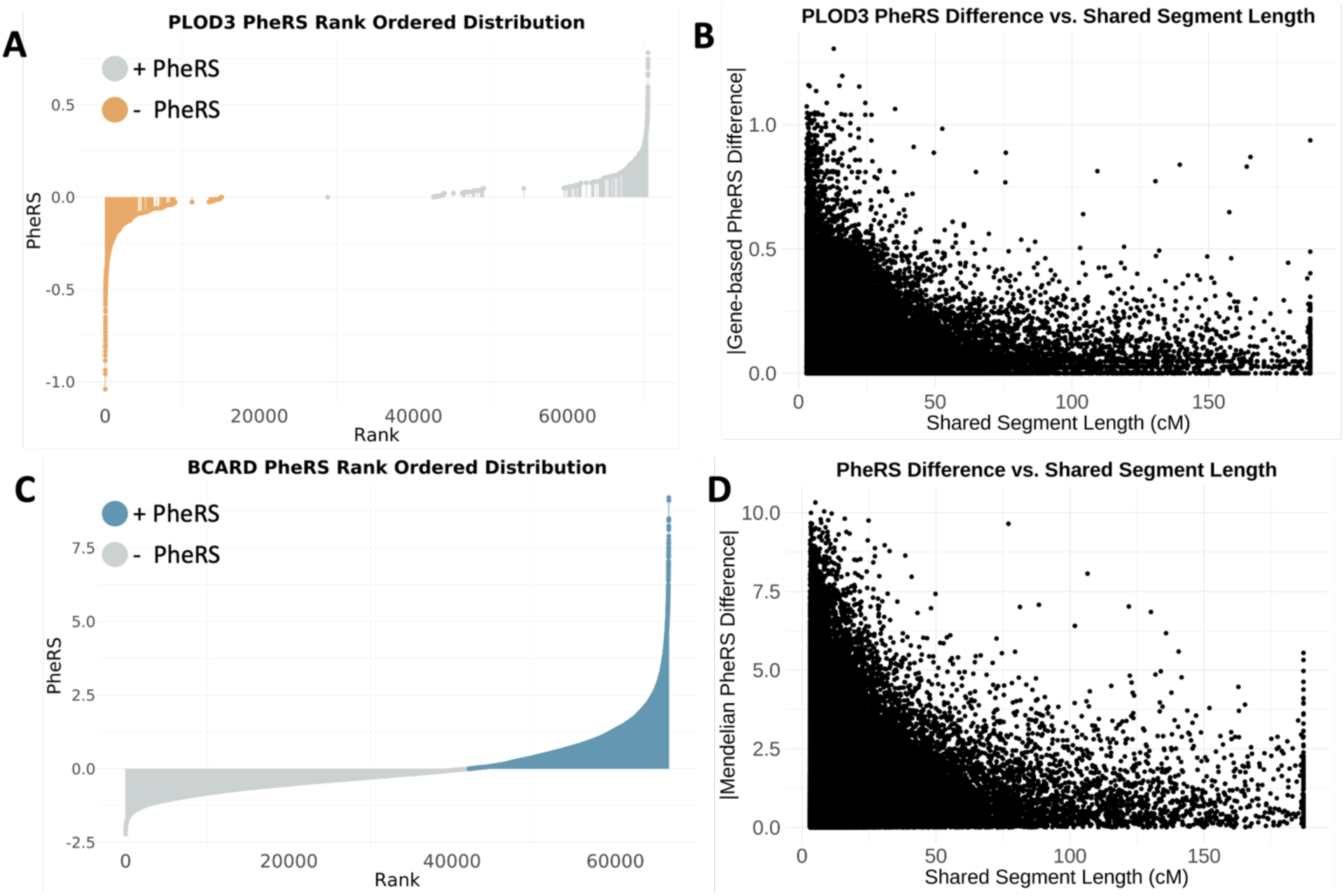
Distribution of Mendelian and gene-based PheRS and the relationship between shared segment length and PheRS. **A)** Plot of the rank ordered distribution of the gene-based PheRS in BioVU. Individuals highlighted in orange with a -PheRS represent those with more of the phenome identified in our gene-based PheWAS. **B)** Plot of pairwise shared segment length by the absolute value of the gene-based PheRS difference. **C)** Plot of the rank ordered distribution of the Mendelian (BCARD) PheRS in BioVU. Individuals highlighted in blue with a + PheRS represent those with more of the BCARD phenome. **D)** Plot of pairwise shared segment length by the absolute value of the Mendelian PheRS difference.

## Notes

### Competing Interest Statement

The authors have declared no competing interest.

### Funding Statement

The Vanderbilt University Medical Center Synthetic Derivative is supported by institutional funding, the 1S10RR025141-01 instrumentation award, and the CTSA grant UL1TR000445 from the National Center for Advancing Translational Sciences/National Institutes of Health. A.S. was supported by the 5T32GM080178-15 and 1T32GM145734-01 from the Training Program in Genetic Variation & Human Phenotypes. M.M.S. was supported by the National Institutes of Health (K12HD043483).

### Author Declarations

The IRB of Vanderbilt University Medical Center gave ethical approval for this work (IRB# 151187).

